# Seasonality and predictors of childhood stunting and wasting in drought-prone areas in Ethiopia: A cohort study

**DOI:** 10.1101/2022.10.09.22280864

**Authors:** Bereket Yohannes Kabalo, Bernt Lindtjørn

## Abstract

**Background and objectives:** Over centuries, Ethiopia has experienced severe famines and periods of serious drought, and malnutrition remains a major public health problem. The aims of this study were to estimate seasonal variations in child stunting and wasting, and identify factors associated with both forms of child malnutrition in drought-prone areas.

**Methods:** This cohort study was conducted among a random sample of 909 children in rural southern Ethiopia. The same children were followed for one year (2017 - 2018) with quarterly repeated measurements of their outcomes: height-for-age and weight-for-height indices (Z-scores). Linear regression models were used to analyse the association between both outcomes and baseline factors (e.g., household participation in a social safety net program and water access) and some time-varying factors (e.g., household food insecurity).

**Results:** Child wasting rates (i.e., low weight-for-heigh Z-scores) varied with seasonal household food insecurity (□^2^ _trend_ = 15.9, *p* = 0.001), but stunting rates (i.e., low height-for-age Z-scores) did not. Household participation in a social safety net program was associated with decreased stunting (*p =* 0.001) and wasting (*p =* 0.002). In addition to its association with decreased wasting (*p =* 0.001), protected drinking water access enhanced the association between household participation in a social safety net program and decreased stunting (*p =* 0.009). Absence of a household latrine (*p =* 0.011), lower maternal education level (*p =* 0.001), larger family size (*p =* 0.004), and lack of non-farming income (*p =* 0.002) were associated with increased child stunting.

**Conclusions:** Seasonal household food insecurity was associated with child undernutrition in rural Ethiopia. Strengthening community-based food security programs, such as the Ethiopian social safety net program, could help to reduce child undernutrition in drought-prone areas. Improving clean water access and sanitation could also decrease child undernutrition.

**Limitations and strengths of this study:**

- This study accounted for agro-ecological strata and random selection of households; however, we did not initially estimate separate samples for the different strata (e.g., by household participation social safety net program).
- Seasonality was accounted for in our repeated measurements using some time-varying exposure variables (e.g., household food insecurity and child diarrhoeal illness); however, we did not assess other illnesses, such as malaria, measles, tuberculosis, etc.
- We did not initially estimate separate samples for our repeated measurements, but our analytical sample (i.e., counts of observations compiled from the repeated measurements) showed adequate statistical power of estimation to meet the primary aims of the study.
- This study analysed height-for-age Z-score and weight-for-height Z-score indices as continuous measures using linear regression models; our estimates were adjusted for the observed clustering effects at the primary sampling stage (at the kebele level) and time series effects of our repeated measurements.
- As our data were analysed in a cohort design, a certain underestimated time element may exist in our repeated measurements beyond the scope of our time-varying exposures.

## INTRODUCTION

Over the centuries, Ethiopia has experienced droughts and famine events, averaging once per decade ^1, 2^. The Intergovernmental Panel on Climate Change (IPCC) also predicts that climate change will lead to more desertification and crop failures in vulnerable areas, such as Ethiopia ^3–5^ where malnutrition remains one of the major public health problems ^6–9^.

In communities that rely on subsistence agriculture, food insecurity often peaks seasonally in pre-harvest periods ^10, 11^. Affected households adapt by: reducing the frequency and size of daily meals; selling livestock or dairy products; borrowing food or money from merchants or local social networks; selling wood, charcoal, or grass; engaging in paid labour; and renting out farm land ^12^. However, such coping mechanisms are often insufficient, leading to a vicious cycle of poverty and food insecurity.

Following many years of food provision for famine-prone populations, the Ethiopian government launched the Productive Safety Net Program (PSNP) in 2005 ^13^. PSNP’s stated aims are to prevent household asset depletion and strengthen communities ^13^. This program provides cash or food payments in exchange for labour to produce public works, local infrastructure, and environmental projects, or direct payments for households that are unable to provide labour ^14^.

Researchers use several methods to assess the impact of food scarcity on populations. One common outcome is the level of household food insecurity and occurrence of malnutrition among vulnerable populations (e.g.., young children); both these outcomes are subjected to the impacts of climate change ^15, 16^. Recent literature has emphasised the importance of understanding seasonal variations in nutrition status among young children as an indicator of vulnerability to food insecurity ^17^. Population’s vulnerability in this context could be described as the level of susceptibility to food insecurity challenges due to lack of capacity to cope and adapt to those challenges, and often affected by several physical, social, economic and environmental factors or processes ^18^.

Since the 1970s, undernutrition has been classified in two major ways: wasting (i.e., low weight-for-height or small mid-upper arm circumference) and stunting (i.e., low height-for-age) ^19^. However, this categorical classification neglects many children with borderline measurements who are undernourished ^20^. As continuous nutritional outcomes, height-for-age and weight-for-height indices (Z-scores) could reflect shared factors and pathophysiological processes that should be viewed as composite measures of undernutrition ^21^.

Wolaita, a densely populated area in southern Ethiopia, has experienced severe droughts and famines ^2, 10^. As such, the population’s experiences and coping mechanisms related to severe food insecurity are relevant for other famine-prone areas in the country and across Africa. The aims of this study were to estimate seasonal variations in child stunting and wasting, and identify factors associated with both these forms of child malnutrition in drought-prone areas.

## METHODS

### Study design and setting

We conducted a prospective cohort study using a random sample of 909 households in the rural Wolaita area in southern Ethiopia. We recruited one child per household at the start of the study (in June 2017), and followed the same children by measuring their outcomes, i.e., height-for-age and weight-for-height indices, every three months for one year (June 2017 to June 2018). Our exposure variables included background factors (measured at baseline) and some time-varying factors (measured each season). Quarterly repeated measurements were performed in the first month of each season (i.e., June, September, December, and March) ^10^.

Wolaita is located between the Great Rift Valley and the Omo Valley in southern Ethiopia. Rural villages in this area mainly represent two agro-ecological areas: the hot and semi-dry ‘lowlands’ and the relatively cooler and sub-humid ‘midlands’ ^3, 22^. Mean annual rainfall ranges from 800 mm in the lowlands to 1,200 mm in the midlands, with a bimodal distribution ^10^. Farming of staple crops, such as maize, occurs during the *Belg* rains from approximately March to early May ^10, 23^. Root crops, such as taro and sweet potato, are farmed in both seasons and help to bridge seasonal gaps in food security ^3, 10^.

### Outcomes

The main outcome measures were height-for-age and weight-for-height indices (Z-scores), measured each season for one year and defined based on the World Health Organization (WHO) 2006 child growth standards ^24^. Stunting and wasting were defined as HAZ (height-for-age Z-scores) and WHZ (weight-for-height Z-scores) of −2 standard deviations below the respective WHO standard median.

### Exposures

Our exposure variables included background factors (baseline data) and some time-varying factors (measured each season). Baseline factors comprised the following: (1) child age and sex; (2) parent age and education; (3) household socio-economic conditions, such as family size, source of income, wealth index, and participation in the food security program (PSNP); (4) household latrine ownership; and (5) drinking water access. We considered household food insecurity, dietary diversity, and child diarrhoeal illness as time-varying exposures, for which we carried out repeated measurements.

### Repeated measurements

Our repeated measurements were performed during the four seasons based on agricultural cycles: *Kiremt* is the sowing season in June, July, and August; *Belg* is the main harvest season in September, October, and November; *Bega* is the post-harvest season in December, January, and February; and *Tsedey* is the dry pre-harvest season in March, April, and May ^10^. These quarterly repeated measurements were carried out at the same time for outcomes and time-varying exposures.

### Participants

A multistage random selection of households was conducted. First, we selected two rural districts, or Woredas, representing the two agro-ecological strata in Wolaita: the Humbo district in the lowlands and Soddo Zuria district in the midland area, with the assumption that household food insecurity would be more prevalent in the lowland areas ^25, 26^. Population density was higher in the midland villages than in the lowland villages in the study area. As such, we selected three kebeles (the smallest administrative unit) from the lowland district and two kebeles from the midland district using the complex samples selection feature in SPSS version 25.0 (IBM). Finally, we selected households with children under five years-old and enrolled one child aged 6 - 59 months per household.

To estimate the sample size, we followed an earlier cohort study assessing seasonal variations in wasting prevalence ^27^. The estimated sample size to estimate differences in prevalence rates of wasting 6.6% and 13%, with a 95% level of confidence and 80% power, was 820 children (OpenEpi software). Our study included 909 children.

### Patient and public involvement

No subject involvement.

### Outcome measurements

Height and weight measurements were performed each season. We trained four data collectors on standard techniques for height and weight measurements. After the training, we validated the consistency of their measurements by recruiting 10 children aged below 5 years from another rural village and having all four data collectors (observers) measure each child’s height twice. The overall measurements showed approximately 92% average internal consistency. These four observers recorded height and weight measurements for the actual study. Height (or recumbent length for children younger than 24 months) was measured to the nearest 0.1 cm using a local wooden length board. Weight was measured to the nearest 0.1 kg using a Seca weight scale (*Seca* GmbH & Co. Kg, Hamburg, Germany).

### Exposure measurements

#### Time-invariant factors (baseline)

Children’s age in months, mothers’ age in years, and highest grade of school completed by both parents were recorded. We also recorded family size (number of household members), source of income (exclusively farming vs. generates other additional income), possession of common household assets, and participation in the food security program (data collectors observed PSNP beneficiary cards during household visits). In addition, we recorded household latrine ownership (yes vs. no) and drinking water access (protected vs. unprotected), and only water piped via public tap was as a protected source ^28^. We used principal component analysis to construct a wealth index based on common household assets: (1) housing material of the roof, interior ceilings, floors, and walls; (2) number of livestock owned by the household; (3) land size in hectares; and (4) possession of common assets, such as a radio, mobile telephone, bed, mattress, kerosene lamp, watch, electric or solar panels, chairs, tables, wooden boxes, donkey carts etc., ^10, 29^.

#### Time-varying factors (measured each season)

The time-varying variable, i.e., household food insecurity (HFI), was measured using nine questions in the Household Food Insecurity Access Scale, which has been validated in the study area ^10^. Household dietary diversity was scored using 24-hour recall measurements. Household members were asked about the 12 common food groups in Ethiopia: (1) cereals and breads; (2) potatoes and other roots or tubers; (3) vegetables; (4) fruits; (5) eggs; (6) dairy products; (7) pulses; (8) fish; (9) meat; (10) oil, fat, or butter; (11) sugar or honey; and (12) other foods or condiments (e.g., coffee, tea, other spices, etc.). The responses for the 12 food-groups were used to generate a scale of food intake diversity, i.e., the household dietary diversity score (HDDS) ^30^. The occurrence of childhood diarrhoeal illness was also assessed, which was defined as the passage of three or more loose or watery stools in the preceding 24-hours ^31, 32^, and was assessed during the two weeks prior to the survey dates ^33^.

We generated two categorical variables from the actual HFI observations in our dataset and the time series of our repeated measurements. Quantified as person-time observations, an ordinal HFI measure (i.e., number of seasons with HFI) was generated as an exposure variable to explore dose-response relationships (e.g., between child wasting and HFI). Quantified also as person-time observations, we generated a multinomial HFI measure summarising incidence rates of HFI by the four seasons (0 = food-secure; 1 = HFI in the sowing season; 2 = HFI in the main harvest season; 3 = HFI in the post-harvest season; and 4 = HFI in the dry pre-harvest season) as an exposure variable in our main analysis (i.e., multivariable models). As household food security and dietary diversity are highly correlated entities, we accounted for HDDS as the null category for HFI multinomial measure (i.e., 0 = food-secure) as an exposure variable in our main analysis. Moreover, child diarrhoeal illness was considered as a covariate for the effect of the other time-varying exposures (e.g., seasonal HFI).

#### Conceptual framework

Based on a systematic review paper, Phalkey and colleagues suggested complex pathways from climate variability to undernutrition in subsistence communities ^34^. Our current work used their work, but we adapted it to the scope of our study, and we focused on human nutrition (Figure 1).

**Figure 1.** Conceptual framework for a possible chain of relationships between seasonal food insecurity and child undernutrition, Wolaita, rural Ethiopia, 2017 - 2018.

## Data and measurements

### Data entry and cleaning

Data were double-entered in EpiData software version 3.1 (EpiData Association 2000 - 2021, Aarhus, Denmark) and corrected for entry errors. First, we entered our baseline data by unique identification numbers for the subjects (ID). We then entered repeated measurements by the subject ID and recorded each round of measurement with different variable names for each variable. After cleaning our data in the short format (by ID), we reshaped the dataset into long format for statistical analysis, with which a new variable (season) was generated to specify the discrete time series of our repeated measurements. We generated nutritional indices (HAZ and WHZ) from anthropometric data using ENA and WHO Anthro software 3.2.2 (WHO, Geneva, Switzerland).

### Units of analysis

As we measured each child in each of the four seasons, we compiled counts of observations totalling 3636 HAZ and WHZ measurements at the end of the study period. However, we excluded 46 HAZ and 126 WHZ observations that had incomplete data or that were severe outliers ^35^. Accordingly, our units of analysis were counts of measurements totalling 3,571 HAZ estimates and 3,510 WHZ estimates (Figure 2). We analysed complete WHZ data (n = 3,510) of 897 children and complete HAZ data (n = 3,571) of 907 children.

**Figure 2.** Flow chart of child anthropometric measurements considered for this cohort study, Wolaita, rural Ethiopia, 2017 - 2018.

### Time-varying data considerations

As we measured the same children in each season, age changes during the study period could lead to certain deviations in outcome estimates (i.e., cohort effects). As such, we generated a separate variable (age in months divided by age in the logarithmic scale) to account for cohort effects. Time-varying effects could also be due to external factors (e.g., seasonality). Accordingly, we considered HFI as a multinomial variable to estimate the seasonally variable effect of household food insecurity on child undernutrition. Furthermore, we accounted for the time series of our repeated measurements using some dummy variables as measurement components of time-varying exposures.

### Statistical methods

We used Stata version 15 (Stata Corp LLC, College Station, TX, U.S.A.) for our statistical works. To explore our data distributions (bivariate analysis), we used parametric tests, such as t-tests to compare two means, ANOVA tests to compare more than two means, and correlation tests to assess the associations between two continuous variables. We analysed our normally distributed data for both outcomes with background factors (baseline data) and some time-varying factors (repeated measurements) using hierarchical linear regression models. Our data comprised two categories: (1) clustering effects at the primary sampling stage (at the kebele level) or (2) time-varying effects within our repeated measurements.

### Multivariable analysis

We first estimated between-variations as main effects of baseline factors on our outcome measures, and then analysed within-variations as main effects to explore time-varying exposure effects on outcome estimates (additional details are provided under separate subheadings hereafter).

### Between-variation models

HAZ data were analysed using a multivariable linear regression model with adjustment for the clustering effect of stunting at the primary sampling stage, but we ignored observed insignificant clustering when analysing the WHZ data ^36, 37^. HAZ and WHZ estimates in the preceding season were considered to control for cohort effects when analysing baseline factors associated with stunting and wasting.

### Within-variation models

At this stage, we aimed to estimate the time-varying exposure effects on outcome estimates, and further analysed the fit between variation models to account for an exposure-season interaction effect (e.g., household food insecurity by the four seasons as a multinomial exposure variable) to estimate seasonally variable effects of relevant exposure variables on outcome estimates. Time-varying exposure effects were estimated as main effects with adjustment for other time-varying effects (i.e., cohort and time series effects) and main effects of all baseline factors included in the fitted models for between-variations.

### Further analysis

We further analysed the fitted models for both outcomes to explore interactions, e.g., additive, or multiplicative effects (e.g., PSNP participation and protected drinking water access on our outcome estimates) or effect modification (e.g., variations in the effect of PSNP participation on child wasting across household food insecurity levels).

### Model reports and meanings

We reported main effects for between- and within-variations using standardized model coefficients (β) with 95% CIs. Decreased model coefficients refer to increased stunting (HAZ) and wasting (WHZ).

## RESULTS

### Participants

Table 1 presents the baseline characteristics of the 907 study participants with complete HAZ data.

**Table 1.**
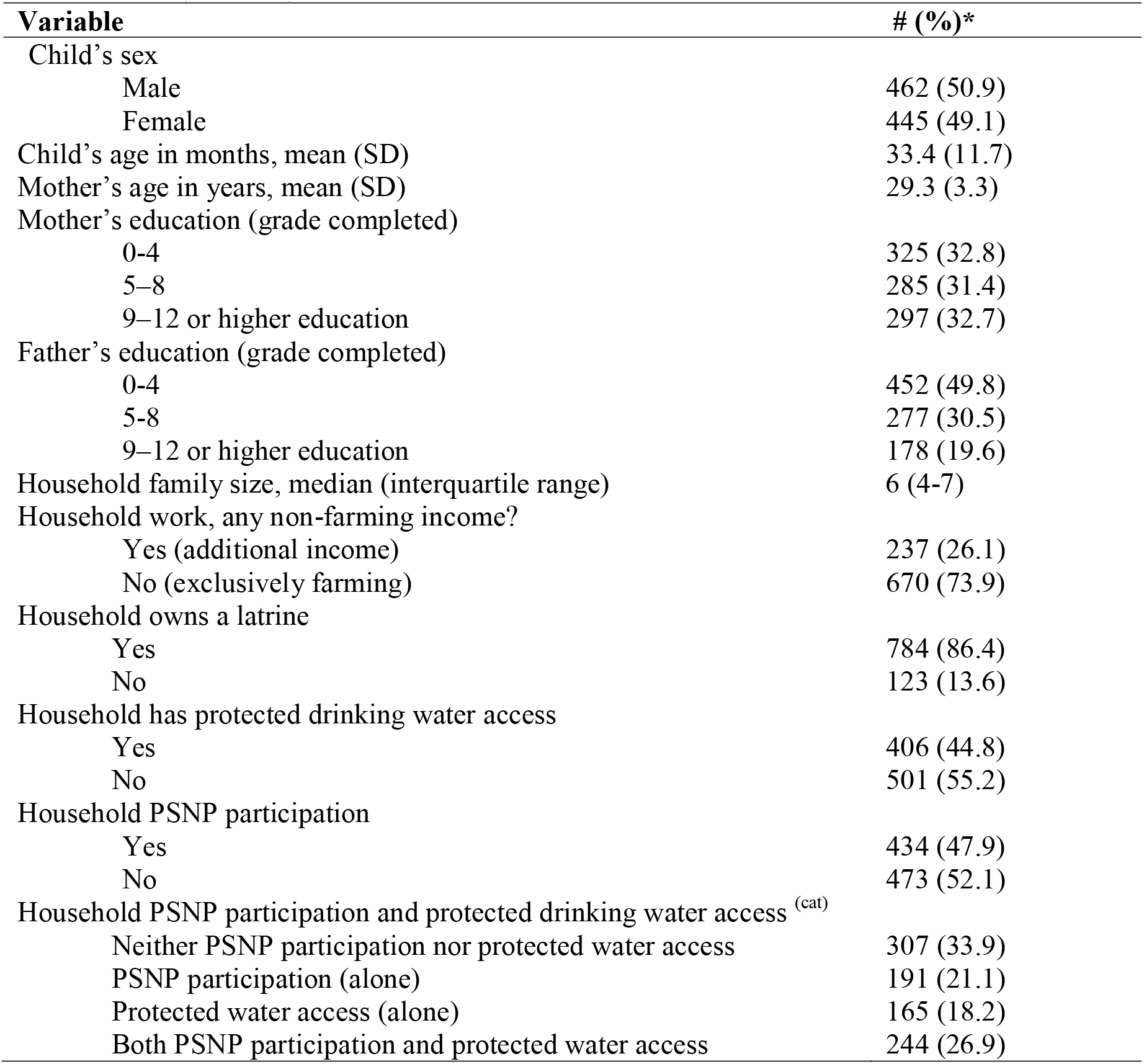
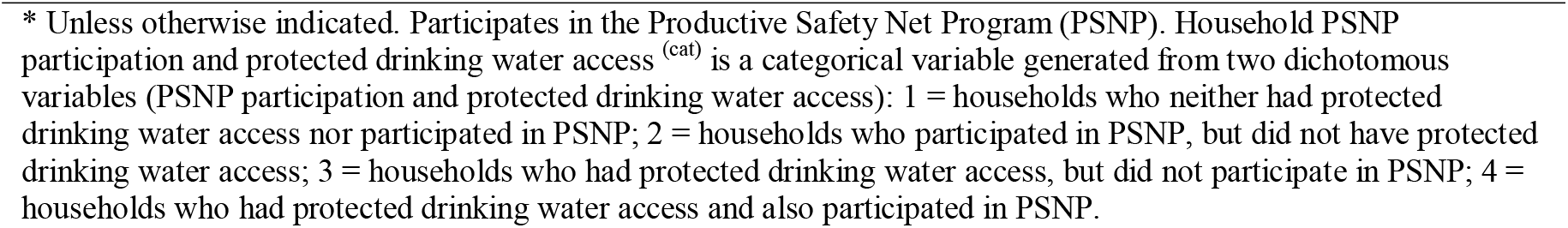
Baseline characteristics of the study participants in rural Wolaita, Ethiopia, 2017 - 2018 (n = 907).

| Variable | # (%)* |
| --- | --- |
| Child's sex |  |
| Male | 462 (50.9) |
| Female | 445 (49.1) |
| Child's age in months, mean (SD) | 33.4 (11.7) |
| Mother's age in years, mean (SD) | 29.3 (3.3) |
| Mother's education (grade completed) |  |
| 0-4 | 325 (32.8) |
| 5-8 | 285 (31.4) |
| 9-12 or higher education | 297 (32.7) |
| Father's education (grade completed) |  |
| 0-4 | 452 (49.8) |
| 5-8 | 277 (30.5) |
| 9-12 or higher education | 178 (19.6) |
| Household family size, median (interquartile range) | 6 (4-7) |
| Household work, any non-farming income? |  |
| Yes (additional income) | 237 (26.1) |
| No (exclusively farming) | 670 (73.9) |
| Household owns a latrine |  |
| Yes | 784 (86.4) |
| No | 123 (13.6) |
| Household has protected drinking water access |  |
| Yes | 406 (44.8) |
| No | 501 (55.2) |
| Household PSNP participation |  |
| Yes | 434 (47.9) |
| No | 473 (52.1) |
| Household PSNP participation and protected drinking water access<sup>(cat)</sup> |  |
| Neither PSNP participation nor protected water access | 307 (33.9) |
| PSNP participation (alone) | 191 (21.1) |
| Protected water access (alone) | 165 (18.2) |
| Both PSNP participation and protected water access | 244 (26.9) |

\* Unless otherwise indicated. Participates in the Productive Safety Net Program (PSNP). Household PSNP participation and protected drinking water access<sup>(cat)</sup> is a categorical variable generated from two dichotomous variables (PSNP participation and protected drinking water access): 1 = households who neither had protected drinking water access nor participated in PSNP; 2 = households who participated in PSNP, but did not have protected drinking water access; 3 = households who had protected drinking water access, but did not participate in PSNP; 4 = households who had protected drinking water access and also participated in PSNP.

### Summary results from repeated measurements

Table 2 summarizes prevalence rates of household food insecurity, childhood diarrhoeal illness, and stunting and wasting. The overall prevalence of child stunting was 32.1% (1,145 of 3,571 measurements) and that of wasting was 10.3% (360 of 3,510 measurements). Child wasting rates varied across seasons (□^2^ _trend_ = 20.5, *p =* 0.001), but stunting rates did not exhibit any seasonal variations.

**Table 2.**
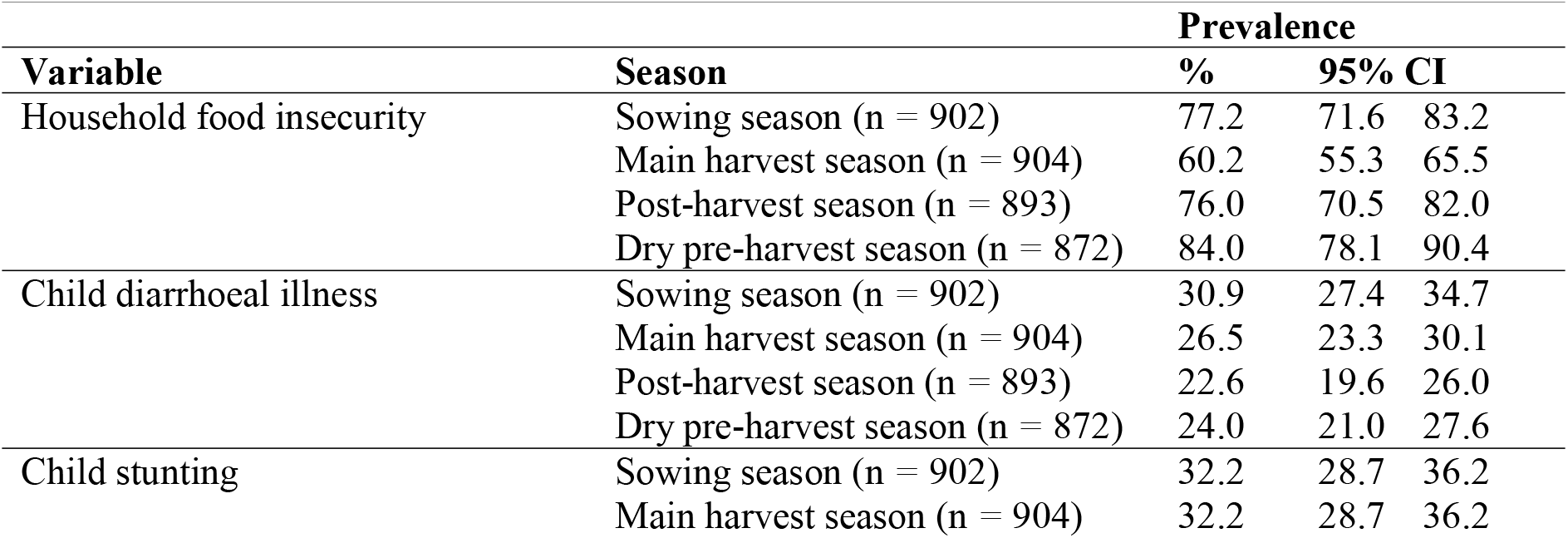
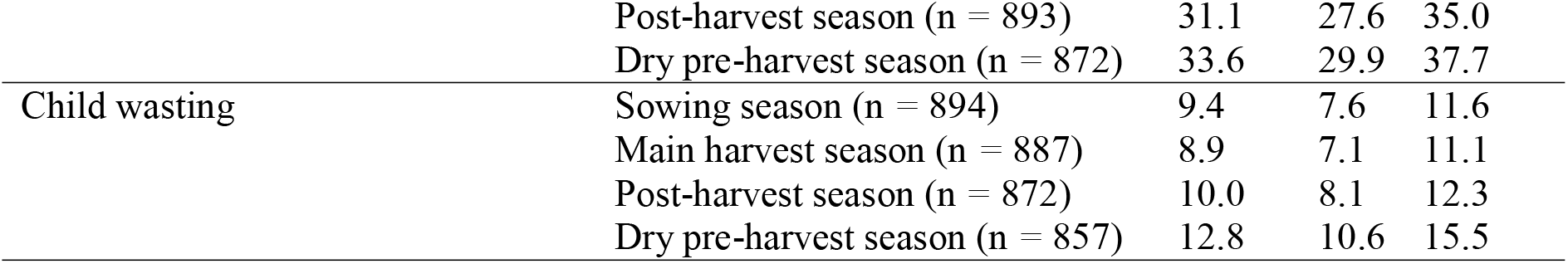
Prevalence rates of household food insecurity, child diarrhoeal illness, and undernutrition (stunting and wasting) rates (Wolaita, rural Ethiopia, 2017 - 2018).

| Variable | Season | Prevalence |  |  |
| --- | --- | --- | --- | --- |
|  |  | % | 95% CI |  |
| Household food insecurity | Sowing season (n = 902) | 77.2 | 71.6 | 83.2 |
|  | Main harvest season (n = 904) | 60.2 | 55.3 | 65.5 |
|  | Post-harvest season (n = 893) | 76.0 | 70.5 | 82.0 |
|  | Dry pre-harvest season (n = 872) | 84.0 | 78.1 | 90.4 |
| Child diarrhoeal illness | Sowing season (n = 902) | 30.9 | 27.4 | 34.7 |
|  | Main harvest season (n = 904) | 26.5 | 23.3 | 30.1 |
|  | Post-harvest season (n = 893) | 22.6 | 19.6 | 26.0 |
|  | Dry pre-harvest season (n = 872) | 24.0 | 21.0 | 27.6 |
| Child stunting | Sowing season (n = 902) | 32.2 | 28.7 | 36.2 |
|  | Main harvest season (n = 904) | 32.2 | 28.7 | 36.2 |
|  | Post-harvest season (n = 893) | 31.1 | 27.6 | 35.0 |
|  | Dry pre-harvest season (n = 872) | 33.6 | 29.9 | 37.7 |
| Child wasting | Sowing season (n = 894) | 9.4 | 7.6 | 11.6 |
|  | Main harvest season (n = 887) | 8.9 | 7.1 | 11.1 |
|  | Post-harvest season (n = 872) | 10.0 | 8.1 | 12.3 |
|  | Dry pre-harvest season (n = 857) | 12.8 | 10.6 | 15.5 |

### Household food insecurity

The overall prevalence of HFI was 74.2% (95% CI 72.8-75.6) during the study period (2,651 of 3,571 measurements), and HFI varied across seasons (□^2^ _trend_ = 29.2, *p =* 0.001). Quantified as person-time observations, the risk of experiencing HFI in just one season was 7.5%, compared to 17.0% in two seasons, 39.0% in three seasons, and 34.8% throughout all four seasons. The median HDDS was 3 (out of the 12 food groups, interquartile range of 3–4), but the overall HDDS did not show seasonal variations. As shown in Figure 3, dietary diversity decreased with increasing durations of household food insecurity (Z = −7.3, *p* = 0.001).

**Figure 3.** Household dietary diversity score (HDDS) by duration of household food insecurity for households who participated in the Productive Safety Net Program (PSNP) and households that did not participate in the program, Wolaita, rural Ethiopia, 2017 - 2018.

### Child diarrhoeal illness

The overall prevalence of child diarrhoeal illness was 26% (95% CI 24.6-27.5; 930 of 3,571 measurements), and the occurrence of child diarrhoeal illness varied across seasons (□^2^ _trend_ = 12.0, *p =* 0.001), with a peak in the rainy (sowing) season.

### Outcome data distributions

Table 3 shows HAZ and WHZ data distributions by background characteristics (baseline data). Boys had lower HAZ indices than girls (t = −4.2, *p =* 0.001). HAZ indices increased (t = 11.6, *p =* 0.001), but WHZ decreased (t = −9.4, *p =* 0.001), with increasing maternal age. Both HAZ and WHZ increased with increasing maternal education level: HAZ (t = 3.8, *p =* 0.001) and WHZ (t = 2.0, *p =* 0.039). Children who lived in exclusively farming households had lower HAZ indices than those who lived in households with additional income (t = −2.9, *p =* 0.004), and WHZ decreased with decreasing household wealth (t = 3.2, *p =* 0.002).

**Table 3.**
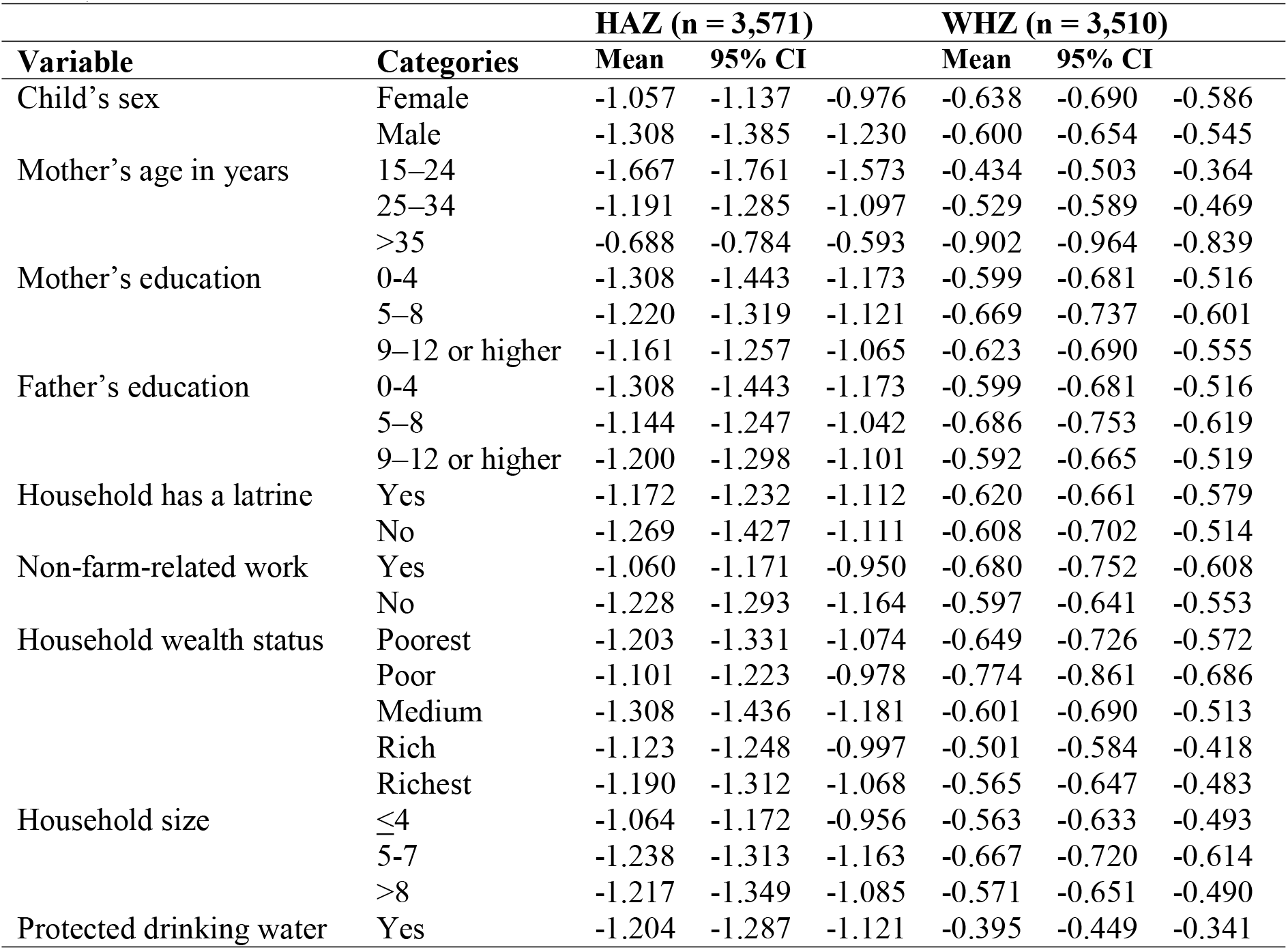
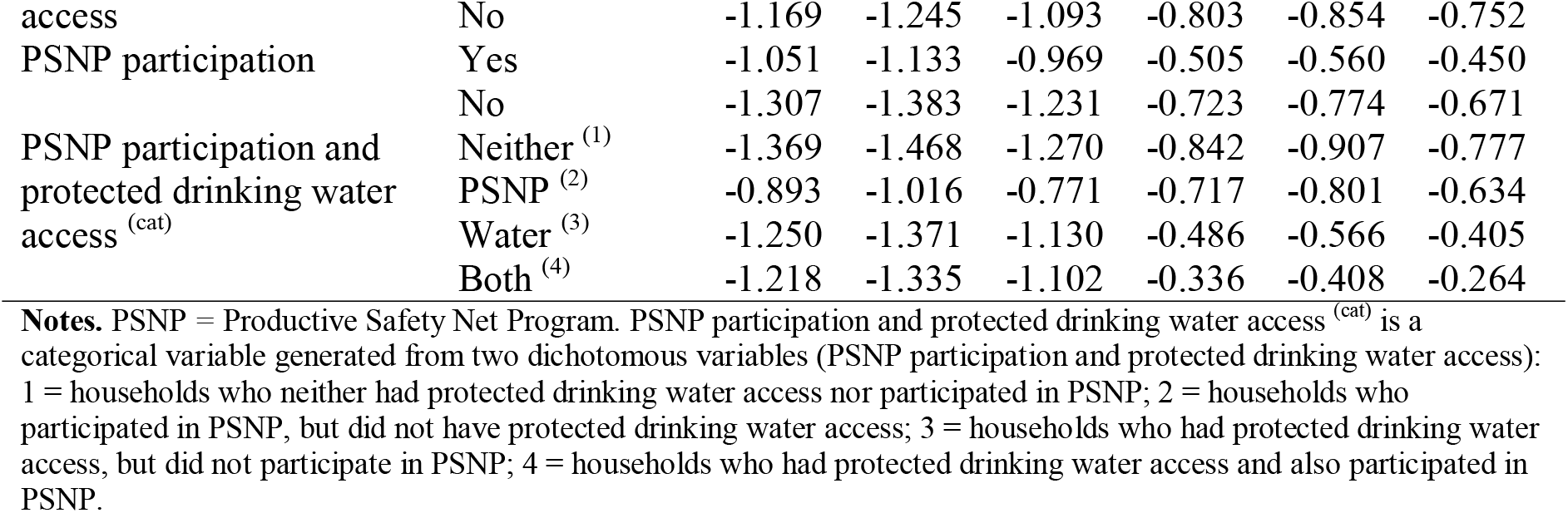
Distributions of height-for-age Z-score (n = 907 children) and weight-for-height Z-score (n = 897 children) indices by baseline factors (rural Wolaita, Ethiopia, 2017 - 2018).

### HAZ and WHZ by PSNP and drinking water access

As shown in Table 3, both HAZ and WHZ indices were higher among children whose households participated in PSNP than those whose households did not participate in PSNP: mean HAZ difference = 0.251 (95% CI 0.136, 0.365) and mean WHZ difference = 0.216 (95% CI 0.141, 0.291). Moreover, children whose households had protected drinking water access had higher WHZ indices than those whose households had no protected drinking water access: mean difference = 0.399 (95% CI 0.324, 0.474). HAZ did not vary by drinking water access.

Protected drinking water access had a confounding effect on the observed association between PSNP participation and increased WHZ: mean difference = 0.136; (95% CI 0.060, 0.212). Figure 4 shows WHZ data distributions by categories for a multinomial variable that disentangles PSNP participation and protected drinking water access.

**Figure 4.** Mean weight-for-height Z-score indices of children aged 6 - 59 months by household participation in the Productive Safety Net Program (PSNP) and protected drinking water access, Wolaita, rural Ethiopia, 2017 - 2018.

### Baseline factors associated with child nutritional status

Table 4 presents results of multivariable regression models for factors independently associated with HAZ and WHZ. PSNP participation was found to be independently associated with increased HAZ and WHZ. In addition, protected drinking water access was associated with increased WHZ. Furthermore, protected water access enhanced the association between PSNP participation and increased HAZ. Being a boy, a lower level of maternal education, and a lack of a household latrine were associated with decreased HAZ.

**Table 4.**
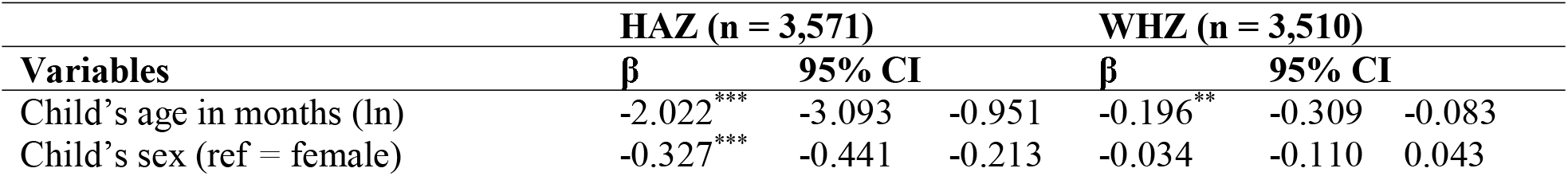
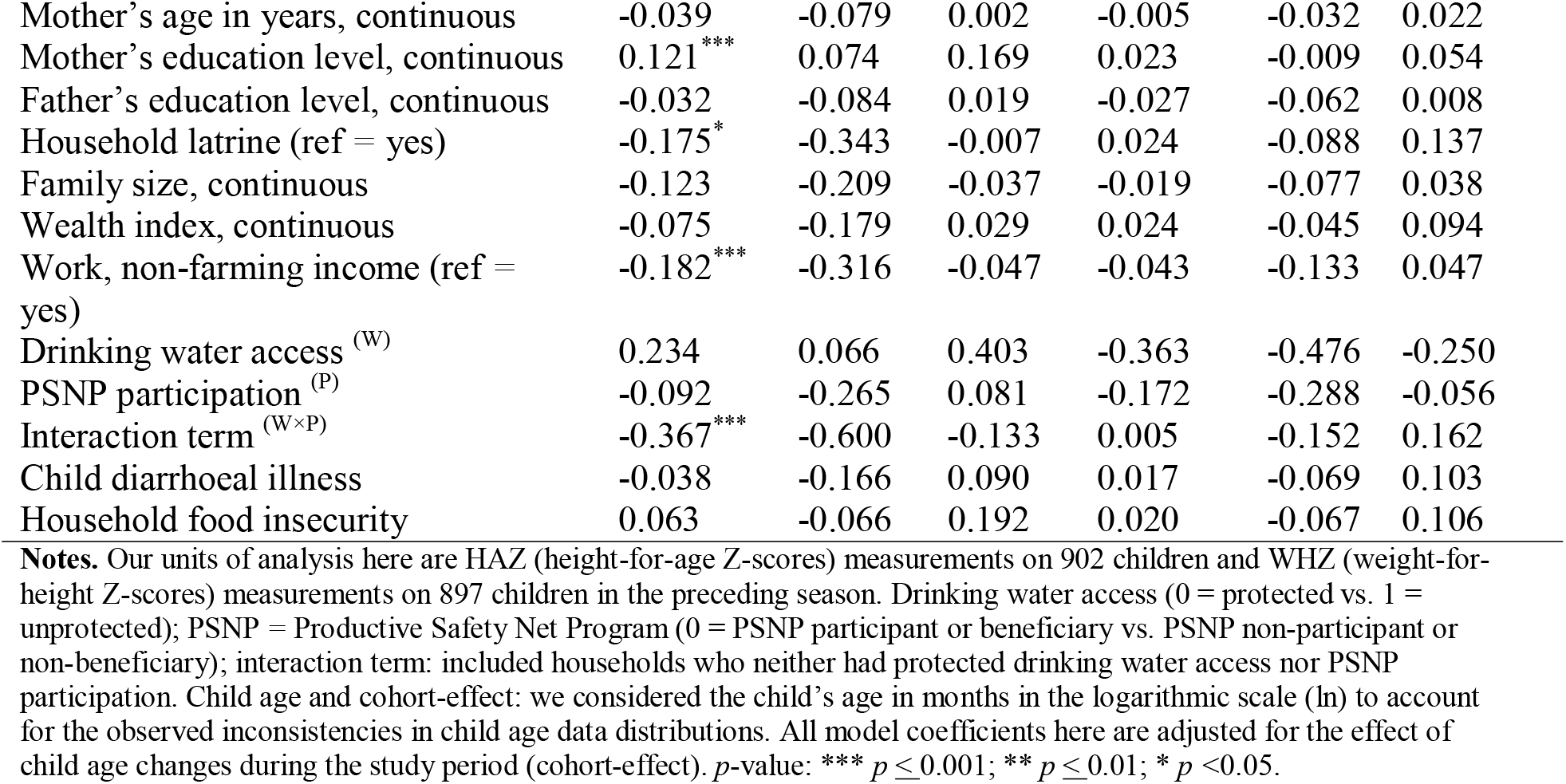
Results of multivariable linear regression models for factors associated with child stunting and wasting (Wolaita, rural Ethiopia, 2017 - 2018).

| Variables | HAZ (n = 3,571) |  |  | WHZ (n = 3,510) |  |  |
| --- | --- | --- | --- | --- | --- | --- |
|  | β | 95% CI |  | β | 95% CI |  |
| Child's age in months (ln) | -2.022*** | -3.093 | -0.951 | -0.196** | -0.309 | -0.083 |
| Child's sex (ref = female) | -0.327*** | -0.441 | -0.213 | -0.034 | -0.110 | 0.043 |
| Mother's age in years, continuous | -0.039 | -0.079 | 0.002 | -0.005 | -0.032 | 0.022 |
| Mother's education level, continuous | 0.121*** | 0.074 | 0.169 | 0.023 | -0.009 | 0.054 |
| Father's education level, continuous | -0.032 | -0.084 | 0.019 | -0.027 | -0.062 | 0.008 |
| Household latrine (ref = yes) | -0.175* | -0.343 | -0.007 | 0.024 | -0.088 | 0.137 |
| Family size, continuous | -0.123 | -0.209 | -0.037 | -0.019 | -0.077 | 0.038 |
| Wealth index, continuous | -0.075 | -0.179 | 0.029 | 0.024 | -0.045 | 0.094 |
| Work, non-farming income (ref = yes) | -0.182*** | -0.316 | -0.047 | -0.043 | -0.133 | 0.047 |
| Drinking water access <sup>(W)</sup> | 0.234 | 0.066 | 0.403 | -0.363 | -0.476 | -0.250 |
| PSNP participation <sup>(P)</sup> | -0.092 | -0.265 | 0.081 | -0.172 | -0.288 | -0.056 |
| Interaction term <sup>(W×P)</sup> | -0.367*** | -0.600 | -0.133 | 0.005 | -0.152 | 0.162 |
| Child diarrhoeal illness | -0.038 | -0.166 | 0.090 | 0.017 | -0.069 | 0.103 |
| Household food insecurity | 0.063 | -0.066 | 0.192 | 0.020 | -0.067 | 0.106 |

**Notes.** Our units of analysis here are HAZ (height-for-age Z-scores) measurements on 902 children and WHZ (weight-for-height Z-scores) measurements on 897 children in the preceding season. Drinking water access (0 = protected vs. 1 = unprotected); PSNP = Productive Safety Net Program (0 = PSNP participant or beneficiary vs. PSNP non-participant or non-beneficiary); interaction term: included households who neither had protected drinking water access nor PSNP participation. Child age and cohort-effect: we considered the child's age in months in the logarithmic scale (ln) to account for the observed inconsistencies in child age data distributions. All model coefficients here are adjusted for the effect of child age changes during the study period (cohort-effect). *p*-value: \*\*\* $p \leq 0.001$ ; \*\* $p \leq 0.01$ ; \* $p < 0.05$ .

### Seasonality

As shown in Figure 5, child wasting rates varied with seasonal household food insecurity, and child wasting rates increased with increasing duration of household food insecurity (□^2^ _trend_ = 5.9, *p =* 0.015). Moreover, Table 5 presents the association between seasonal household food insecurity and child WHZ indices adjusted for baseline factors, child age changes during the study period, and time series of our repeated measurements. It was found that seasonal household food insecurity was independently associated with decreased WHZ.

**Table 5.** Results of multivariable linear regression models for seasonality of child wasting, Wolaita, rural Ethiopia, 2017 - 2018 (n = 3,510).

| Variables | $\beta$ | 95% CI | |
| --- | --- | --- | --- |
| Child's age in months | -0.150** | -0.263 | -0.036 |
| Child's sex (ref = female) | -0.009 | -0.083 | 0.065 |
| Mother's age, continuous | 0.013 | -0.019 | 0.045 |
| Mother's education, continuous | 0.021 | -0.010 | 0.052 |
| Father's education, continuous | -0.021 | -0.054 | 0.013 |
| Household latrine (ref = yes) | 0.050 | -0.059 | 0.159 |
| Family size, continuous | -0.026 | -0.082 | 0.030 |
| Wealth index, continuous | 0.020 | -0.048 | 0.087 |
| Work, non-farming income (ref = yes) | -0.033 | -0.120 | 0.055 |
| PSNP participation | -0.122** | -0.199 | -0.044 |
| Drinking water access | -0.349*** | -0.431 | -0.267 |
| Child diarrhoeal illness (ref = yes) | 0.006 | -0.081 | 0.093 |
| Household food insecurity (ref = yes) | 0.077 | -0.058 | 0.211 |
| Seasonal household food insecurity (ref = main harvest season) |  |  |  |
| Sowing season | 0.003 | -0.131 | 0.137 |
| Post-harvest season | -0.170* | -0.302 | -0.037 |
| Dry pre-harvest season | -0.345*** | -0.481 | -0.209 |
| Child age changes during the study period (cohort) | 0.638* | 0.066 | 1.211 |
| Child diarrhoeal illness (time series) |  |  |  |
| At first season | -0.020 | -0.133 | 0.092 |
| At second season | 0.004 | -0.104 | 0.112 |
| At third season | -0.157 | -0.279 | -0.034 |
| At fourth season | 0.054 | -0.053 | 0.161 |
| Household food insecurity (time series) |  |  |  |
| At first season | 0.167 | 0.071 | 0.263 |
| At second season | -0.143 | -0.225 | -0.061 |
| At third season | 0.192 | 0.052 | 0.333 |
| At fourth season | -0.083 | -0.246 | 0.079 |

*p*-value: \*\*\* $p \leq 0.001$ ; \*\* $p \leq 0.01$ ; \* $p < 0.05$ .

**Figure 5.** Seasonal variations in child wasting rates among overall measurements and measurements within food-insecure households, Wolaita, rural Ethiopia, 2017 - 2018.

### Effect modification

Table 6 presents the results of further analysis of the fitted WHZ model by duration of household food insecurity. The results showed that seasonal household food insecurity modified the effect of PSNP participation and protected drinking water access on increased WHZ estimates.

## DISCUSSION

### Key results

Our cohort study suggested seasonal variations in weight-for-height Z-score (WHZ) indices among children aged 6 - 59 months in rural southern Ethiopia, with dose-response relationships. Household participation in the Ethiopian government’s social safety net (PSNP) and having protected drinking water access were main factors associated with increased height-for-age Z-score (HAZ) or WHZ. Lower level of maternal education, lack of non-farming household income, and absence of a household latrine were associated with decreased HAZ indices.

### Limitations and strengths

This study was based on a random sample of households, and our units of analysis were counts of total observations compiled from four repeated measurements undertaken for one year. However, our one sample estimation could constitute a limitation of this study, especially as certain background factors (e.g., household wealth, PSNP participation, and drinking water access) clustered in the lowland areas. Subjectivity bias could also exist in our food insecurity measurements, especially as this study was conducted in a chronically food-insecure setting ^38, 39^. We used the Household Food Insecurity Access Scale questionnaire, which had been validated in the study area ^10^. To ensure accuracy of the current study data, measurements were repeated and, although we used the same data collectors for the repeated data rounds, it was sometimes not possible to maintain the same data collectors. We did not initially estimate separate samples for our repeated measurements, but our analytical sample (i.e., counts of observations compiled from the repeated measurements) showed adequate statistical power. We analysed HAZ and WHZ indices as composite measures of child nutritional status as continuous variables using linear regression models which enhanced the accuracy of our estimates ^21^. Moreover, we accounted and adjusted our estimates for the observed clustering effects at the primary sampling stage (at the kebele level). We accounted for seasonality through some time-varying exposures (household food insecurity and child diarrhoeal illness), but we did not assess other illnesses, such as malaria, measles, tuberculosis, etc. ^40–43^. We considered some dummy variables to account for the time series random effects beyond the scope of our time-varying exposures.

### Comparative discussions

Our prevalence estimates for stunting and wasting rates are consistent with recent estimates from Ethiopian demographic and health surveys ^44^. Our study indicated that household socioeconomic conditions, such as low level of maternal education, lack of non-farming income, and absence of PSNP participation were associated with HAZ or WHZ indices (Table 4), which complements previous reports ^32, 45–47^. The country’s persistently low socio-economic status is often described as a main cause for child undernutrition in Ethiopia ^32, 45–47^.

Our study suggested that household PSNP participation could improve HAZ and WHZ (Table 4), which complements some extant literature ^48, 49^, but contradictory reports also exist ^50–52^. These inconsistencies could be due to variations in the study designs, participants, and data approaches. Nonetheless, a growing body of literature indicates that PSNP improves household food security and consumption.

Our study points to the lack of clean water access and a household latrine as independent predictors for child undernutrition (Table 4), which aligns with previous reports ^6, 53^. The lack of clean water access and sanitation could predispose populations to recurrent infections, such as diarrhoea and intestinal parasites ^54, 55^. Indeed, improving clean water, sanitation, and hygiene are frequently identified as priority interventions to reduce child undernutrition ^56^.

Our study suggested seasonal household food insecurity as an independent predictor of decreased WHZ (Table 5), which is in accordance with previous investigations ^57, 58^. This could be attributable to seasonal fluctuations in household food consumption patterns ^11, 59^, and some scholars describe seasonality as a grossly neglected dimension of poverty ^11, 17, 60^.

### Meanings and possible explanations

Seasonal household food insecurity could contribute to decreased WHZ indices in rural Ethiopia, suggesting that this population is vulnerable to food insecurity ^17^. Strengthening the PSNP intervention through effective targeting of the neediest households might further enhance its impacts on child undernutrition. Yet, the criteria for household PSNP eligibility remain controversial ^61^; for example, if geographic criteria prioritise the lowland areas ^13^, then poorer households in the midland villages (as in our study setting) could be neglected. Improving clean water access and sanitation have widely been described as nutrition-sensitive interventions ^62, 63^. In our study area context, expanding the coverage of protected drinking water access in the midland villages might contribute to reducing child undernutrition. Moreover, addressing seasonal variations in household food insecurity could help decrease child undernutrition ^17^.

## Data Availability

All data relevant to the study are included in the article or uploaded as supplementary information.

## DECLARATIONS

### Ethics approval and consent to participate

The Institutional Review Committee at Hawassa University in Ethiopia (IRB/002/09) and the Regional Committee for Medical and Research Ethics in Western Norway (2016/482/REK vest) approved the study protocol. Written consent was obtained from the respondents, as well as assent for their children’s weight and height measures. Responses were anonymised using unique identification numbers.

### Consent for publication

Not applicable.

### Competing interests

The authors declare that they have no competing interests.

### Funding

This study was funded by the South Ethiopia Network of Universities in Public Health project, under the Norwegian Program for Capacity Development in Higher Education and Research for Development (Grant Number: ETH-13/0025 “SENUPH”). The funder had no role in associated conduct, writing, or decisions to publish this manuscript.

## Acknowledgements

The authors express gratitude to the Centre for International Health at the University of Bergen in Norway and Hawassa University in Ethiopia for providing facilities to conduct this study. Dr. Eskindir Loha is sincerely acknowledged for his important contribution in carrying out this study. Our field supervisors and enumerators are deeply thanked for their invaluable commitment. The authors gratefully acknowledge the study respondents for their cooperation in repeated rounds of interviews.

## Authors’ contributions

BYK conceived the research idea, designed the study protocol, implemented the study, made major statistical analyses, and wrote the draft and final versions of the scientific report. BL conceived the research idea, and made substantial contributions to the design, field methods, major statistical analyses, and writing. Both authors read and approved this manuscript for submission.

## Notes

### Competing Interest Statement

The authors have declared no competing interest.

### Author Declarations

The Institutional Review Committee at Hawassa University in Ethiopia and the Regional Committee for Medical and Research Ethics in Western Norway approved the study protocol.

