## Supplementary material for "Seasonality and predictors of childhood stunting and wasting in drought-prone areas in Ethiopia: A cohort study": Figure 1

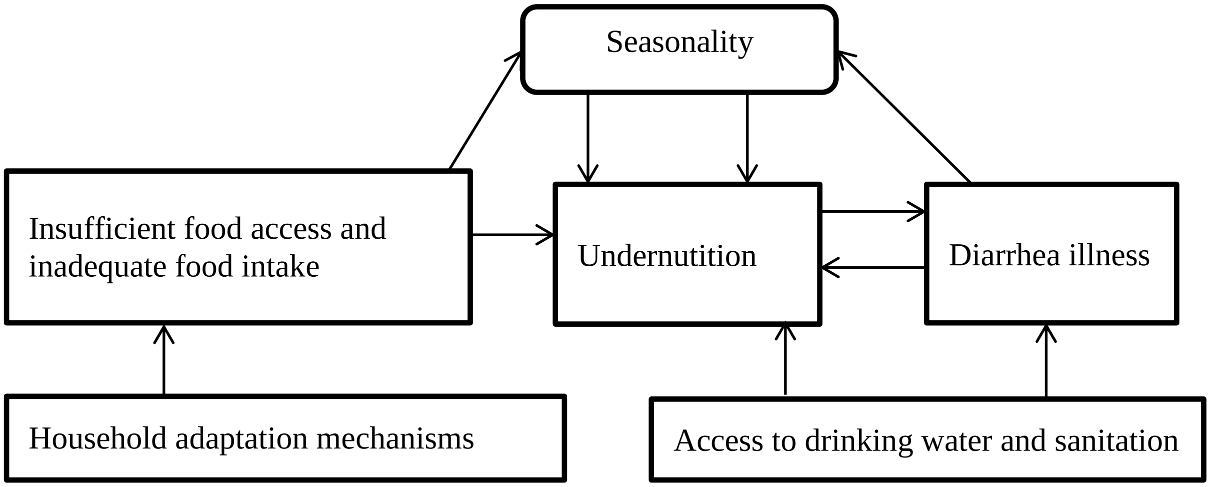


Figure 1 Conceptual framework for possible chain of relationships between seasonal food insecurity and risk for child undernutrition, Wolaita, Ethiopia, 2017-18
