## Supplementary material for "Seasonality and predictors of childhood stunting and wasting in drought-prone areas in Ethiopia: A cohort study": Figure 2

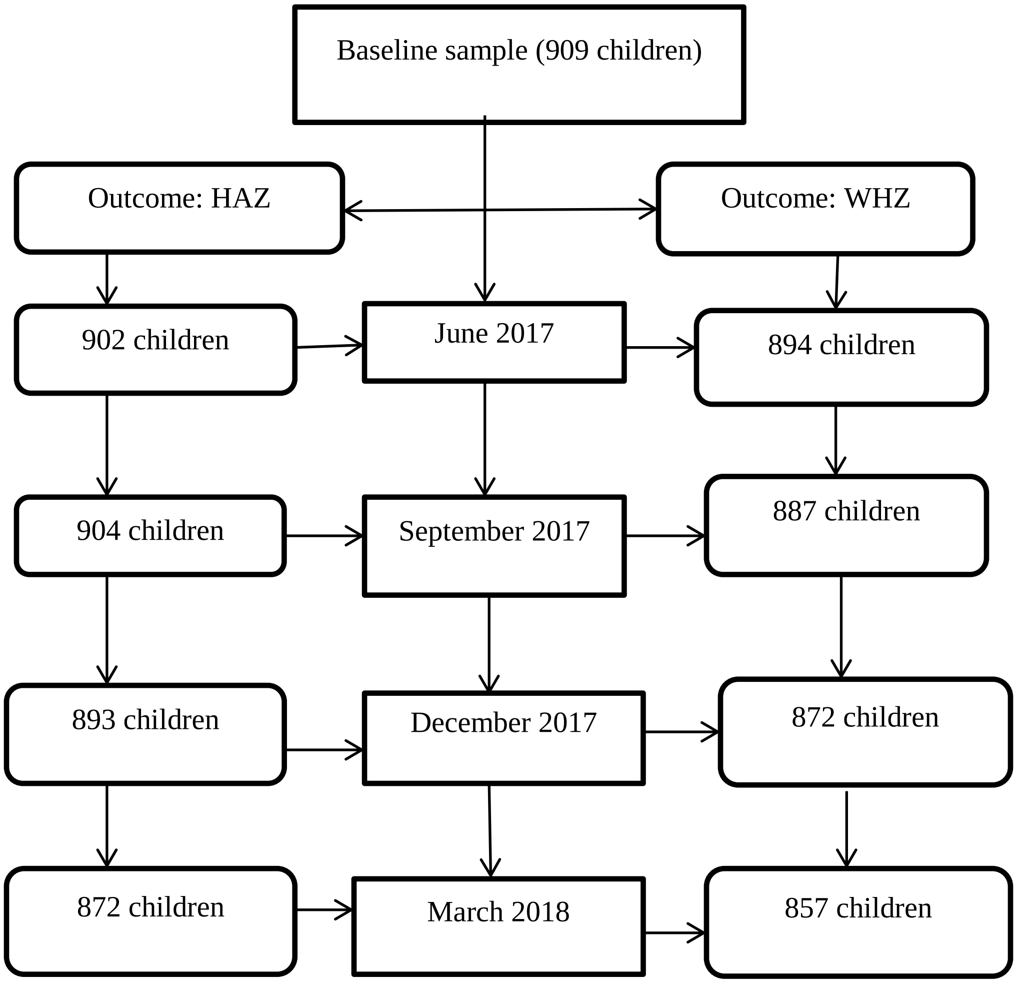


Figure 2 Height-for-age Z-scores (HAZ) and Weight-for-height Z-scores (WHZ) of children aged 6-59 months considered for this cohort study, Wolaita, rural Ethiopia, 2017-18
