## Supplementary material for "Seasonality and predictors of childhood stunting and wasting in drought-prone areas in Ethiopia: A cohort study": Figure 3

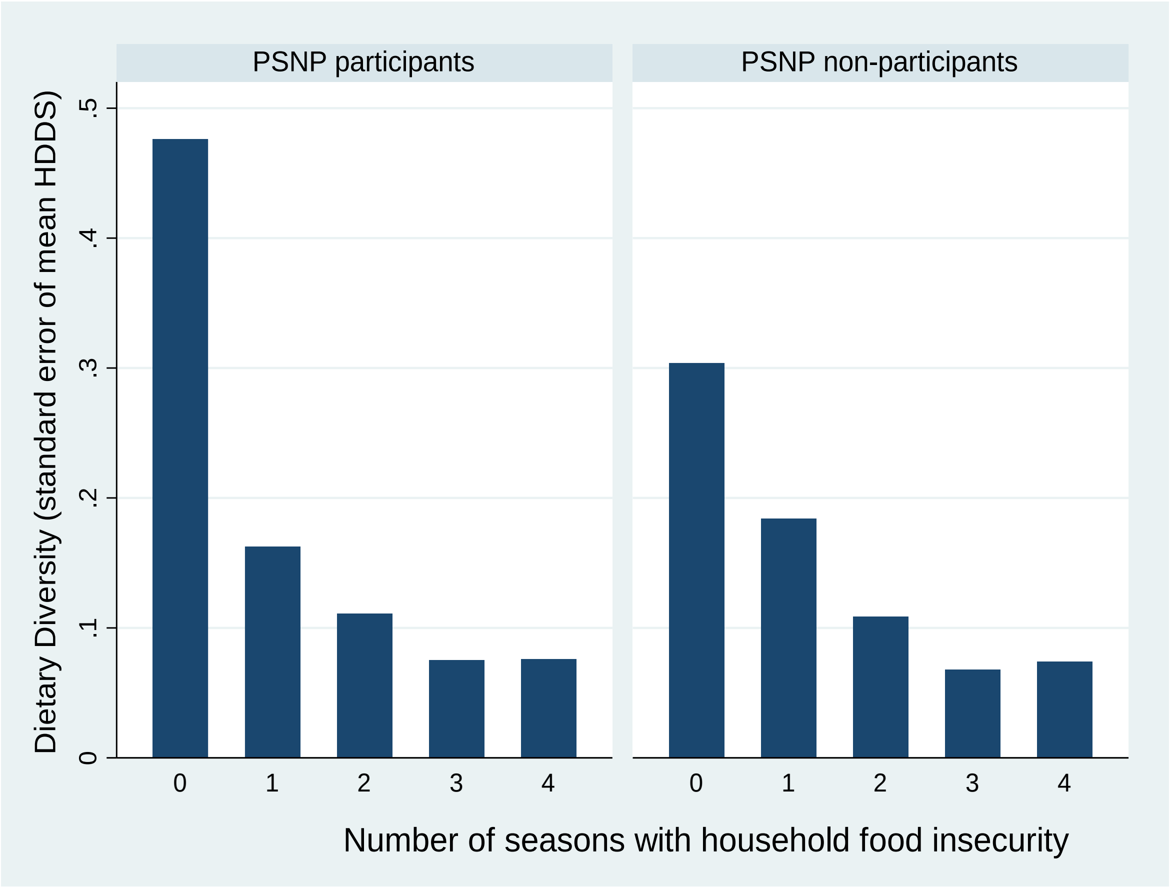


Figure 3 household dietary diversity score (HDDS) by duration of household food insecurity for households who participated in Productive Safety Net Programme (PSNP) and households that did not participate in this program, Wolaita, rural Ethiopia, 2017-18
