## Supplementary material for "Seasonality and predictors of childhood stunting and wasting in drought-prone areas in Ethiopia: A cohort study": Figure 4

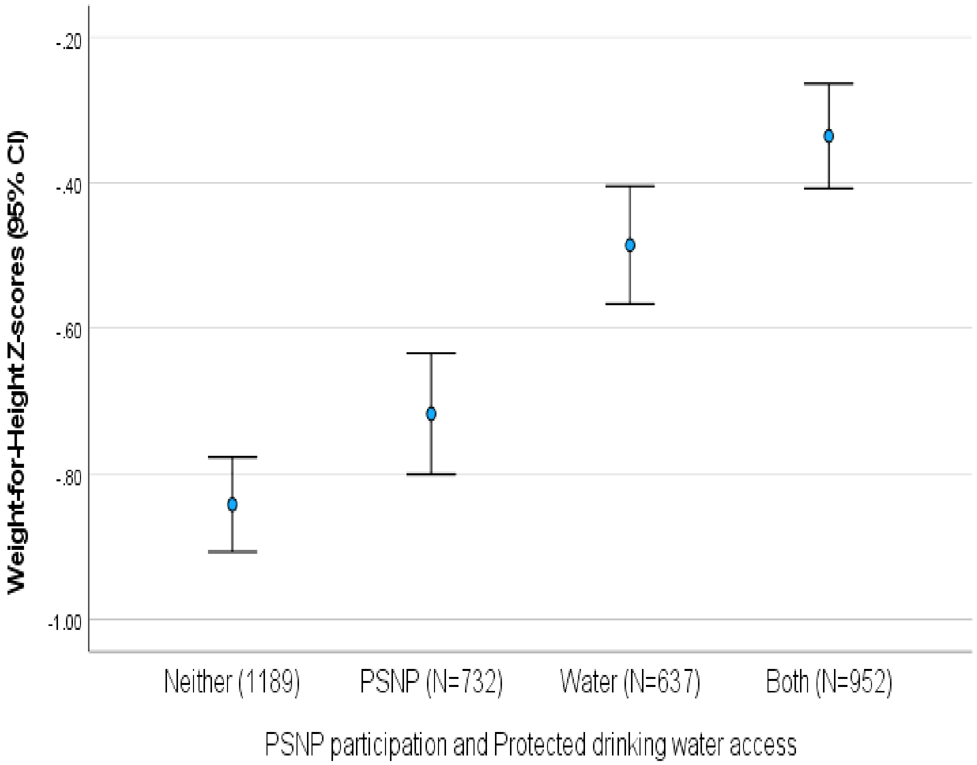


**Figure 4 Weight-for-height Z-scores of children aged 6-59 months by household participation in 'Productive Safety Net Programme (PSNP)' and protected water access, Wolaita, rural Ethiopia, 2017-18**
