## Supplementary material for "Seasonality and predictors of childhood stunting and wasting in drought-prone areas in Ethiopia: A cohort study": Figure 5

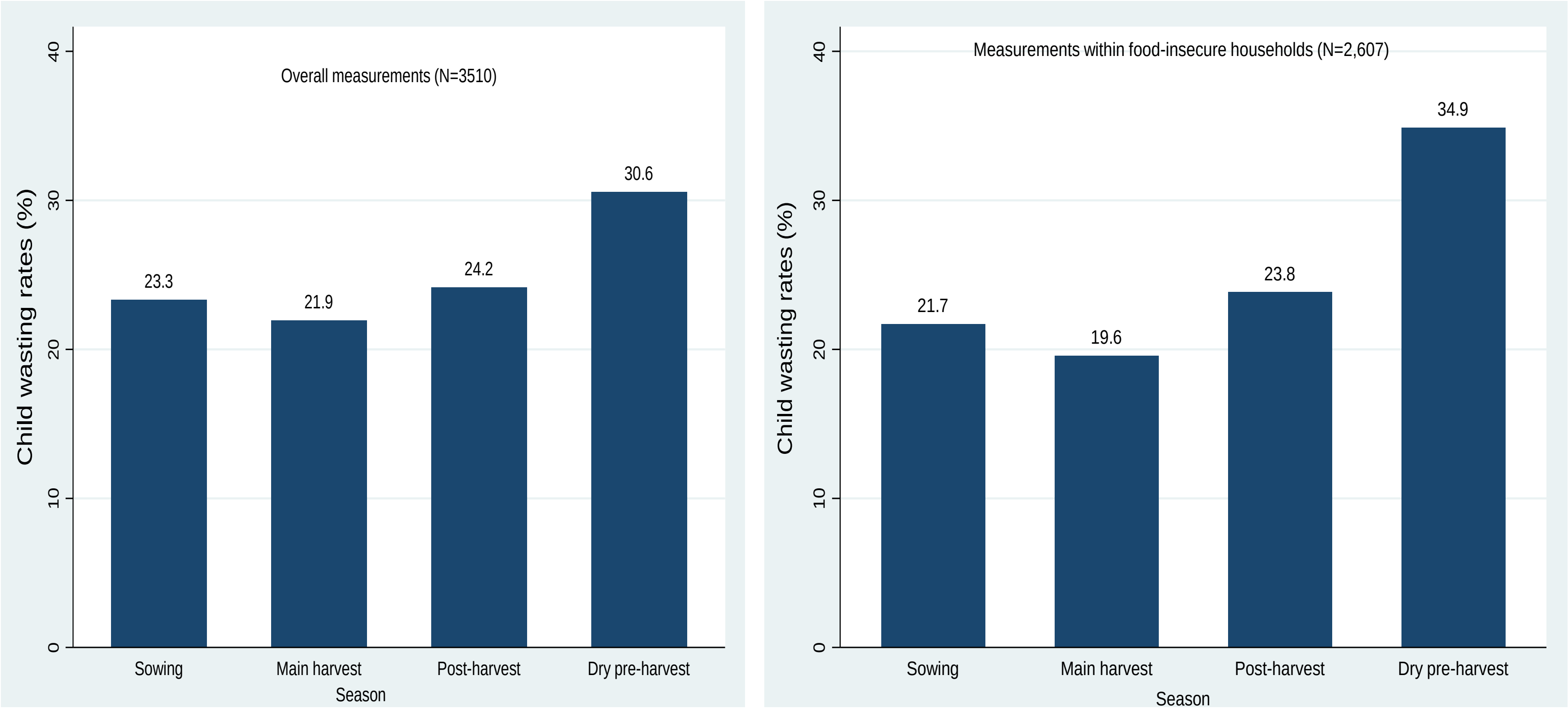


Figure 5 seasonal variations in child wasting rates among overall measurements and measurements within food-insecure households, Wolaita, rural Ethiopia, 2017-18
