## Supplementary material for "Seasonality and predictors of childhood stunting and wasting in drought-prone areas in Ethiopia: A cohort study": Table 6

Table 6 variations in the effects of household PSNP participation and protected drinking water access on weight-for-height Z-scores of children aged 6-59 months by durations of household food insecurity, Wolaita, rural Ethiopia, 2017-18

| | Duration of household food insecurity | | | | | |
|---|---|---|---|---|---|---|
| | 1or 2 seasons | | | 3 or 4 seasons | | |
| | β | 95% CI | | β | 95% CI | |
| Child age (lnmonths), continuous | 1.279 | -0.039 | 2.598 | 1.417 | 0.596 | 2.238 |
| Child age changes (cmonths), continuous | -0.266 | -0.461 | -0.070 | -0.242 | -0.361 | -0.124 |
| Child sex (ref=female) | -0.141 | -0.277 | -0.004 | 0.031 | -0.056 | 0.119 |
| Mother’s age ,continuous | -0.046 | -0.097 | 0.005 | -0.034 | -0.065 | -0.004 |
| Mother’s education, continuous | -0.141 | -0.198 | -0.083 | 0.068 | 0.032 | 0.105 |
| Father’s education ,continuous | 0.156 | 0.092 | 0.220 | -0.080 | -0.120 | -0.041 |
| Household latrine (ref=yes) | -0.382 | -0.592 | -0.171 | 0.178 | 0.049 | 0.307 |
| Family size ,continuous | -0.004 | -0.112 | 0.105 | 0.004 | -0.062 | 0.070 |
| Wealth index, continuous | -0.099 | -0.227 | 0.030 | 0.061 | -0.018 | 0.141 |
| Work, non-farming income | -0.273 | -0.438 | -0.108 | 0.038 | -0.065 | 0.142 |
| PSNP participation and protected drinking water access (cat) (ref=both) | | | | | | |
| Neither (1) | -0.679 | -0.867 | -0.491 | -0.395 | -0.518 | -0.273 |
| PSNP (2) | -0.350 | -0.556 | -0.143 | -0.296 | -0.430 | -0.162 |
| Water (3) | -0.421 | -0.616 | -0.225 | -0.058 | -0.194 | 0.079 |
| Both (4) | | | | | | |
| Child diarrhoeal illness (ref=no) | 0.015 | -0.144 | 0.173 | 0.008 | -0.094 | 0.110 |
| Child diarrhoeal illness (1st season) | -0.066 | -0.275 | 0.142 | 0.088 | -0.046 | 0.222 |
| Child diarrhoeal illness (2nd season) | 0.077 | -0.126 | 0.280 | 0.000 | -0.127 | 0.127 |
| Child diarrhoeal illness (3rd season) | -0.010 | -0.232 | 0.211 | -0.150 | -0.296 | -0.003 |
| Child diarrhoeal illness (4th season) | -0.088 | -0.286 | 0.110 | 0.143 | 0.016 | 0.270 |

Notes. PSNP= Productive Safety Net Programme. PSNP participation and protected drinking water access (cat) is a categorical variable generated from two dichotomous variables (PSNP participation and protected drinking water access): 1=included households who neither had protected drinking water access nor participated in PSNP; 2= households who participated in PSNP but did not have protected drinking water access; 3= households who had protected drinking water access but that did not participate in PSNP but did not have protected drinking water access; 4= households who had both protected drinking water access and PSNP participation. Decreased (β) coefficients refer to increased child wasting and vice-versa. Child age changes (cmonths) =child age in months divided by child age in the logarithmic scale; cmonths was used to account and adjust for child age changes during the study period.
